# Respiratory viral infections do not increase risk of venous thromboembolism

**DOI:** 10.1101/2023.10.02.23296457

**Authors:** Steven J. Korzeniewski, Samantha J. Bauer, Christopher Kabrhel, D. Mark Courtney, Ngai Ka ming, Christopher Kelly, Carlos A. Camargo, Jeffrey A. Kline

## Abstract

**Background:** A major concern in emergency departments is whether acute respiratory illness (ARI) is associated with increased risk of venous thromboembolism (VTE).

**Methods:** This prospective cohort study includes ARI patients from the 91-hospital, U.S. CDC-sponsored Respiratory Virus Laboratory Emergency Department Network Surveillance (RESP-LENS) program from January 2022 to June 2023. We calculated incidence rates and used multivariable regression models to test the null hypothesis that there is no association between the results or absence of laboratory viral testing and the risk of new onset VTE within 30-days.

**Findings:** Out of 620,303 ARI encounters, 65% underwent laboratory viral testing; 13% tested positive for COVID and 3%-4% tested positive for influenza-A and/or RSV. The 30-day VTE incidence rate was 0.**70% among** unique first patient encounters and 0.**82%** overall.

The highest VTE incidence rate occurred among viral test negative patient encounters (1.13% [95%CI 1.08%-1.19%]) and the lowest VTE rate occurred among patients who tested positive for COVID, influenza-A or RSV, or who did not undergo viral testing (95%CI 0.09%-0.86%). In adjusted models, only among patients receiving any ICU care was fluA associated with heightened VTE risk.

New VTE was associated with increased 30-day mortality risk (RR 2.7, 95%CI 2.3-3.2), but there was no difference in 30-day mortality risk among VTE patients grouped by the results of viral testing (interaction p > 0.05).

**Conclusion:** In the U.S. from January 2022 to June 2023, laboratory confirmed viral infection was not associated with increased risk of short-term VTE diagnosis or death among patients seeking emergency care.

## INTRODUCTION

In the United States (U.S.), at least 120 million people visit an emergency department (ED) each year, and about 5-10% of them present with acute respiratory illnesses (ARI) that include viral infections.^1^ A major concern is whether ED patients with positive viral tests are at increased risk of developing venous thromboembolism (VTE). Well-established thrombogenic viruses include hepatitis B and C, HIV and cytomegalovirus,^2^ but less is known about respiratory viruses.

Most of the information about an association between influenza A (fluA) and VTE is specific to the H1N1 subtype.^3,4^ Despite its increased prevalence among infants and the elderly, we were unable to identify a previous report of an association between respiratory syncytial viral (RSV) and VTE. While SARS-CoV-2 is widely characterized as thrombogenic, VTE risk estimates range anywhere from 0%-to-∼40% in different clinical settings, and most of the published data are specific to a period of Hunan strain predominance in patients with severe infection.^5-9^ By contrast, members of our group conducted a large network study of ambulatory patients that found a decreased risk of VTE among symptomatic laboratory confirmed COVID positive patients compared to laboratory confirmed test negative patient encounters.^10^

In this manuscript, we leverage novel data from a large national ED surveillance network that was established in late 2021 with support from the U.S. Centers for Disease Control and Prevention (CDC). We investigated whether the incidence of VTE within 30-days is increased among encounters with laboratory confirmed viral test positive ARI patients, as well as testing for differences in mortality risk associated with VTE among ARI patients grouped by the results (+/-) or absence of viral testing.

## METHODS

### Study design

This prospective cohort study includes information collected by the Respiratory Virus Laboratory Emergency Department Network Surveillance (RESP-LENS) program from January 2022 to June 2023 to represent all seasons.^11^. Additional details of the methods used by RESP-LENS are *in press* elsewhere and were made publicly available as a preprint 1(DOI: 0.13140/RG.2.2.16885.55521).^12^ Briefly, RESP-LENS involves the collaboration of 24 investigators representing 91 hospitals and the CDC who prospectively surveil viral infections (https://www.cdc.gov/surveillance/resp-lens/dashboard.html). The network began in September 2021, and 22 of 24 sites, covering all ten U.S.

Department of Health and Human Services regions, were reporting data as of January 2022 (Supplemental Table 1). The protocol was reviewed by all site institutional review boards and was either deemed non-human subjects research (i.e., surveillance), exempted from review or approved with expedited review. Importantly, RESP-LENS adheres to the STROBE (Strengthening the Reporting of Observational Studies in Epidemiology) and RECORD (Reporting of studies Conducted using Observational Routinely-collected Data) reporting guidelines.^13,14^

### Procedures

We used the DMP tool to make the RESP-LENS data management plan publicly available at https://dmptool.org/plans/70148. Data were collected for ARI patients at two times; first, from the qualifying index visit and then follow-up data were collected 30 days later. If the patient was discharged on the index visit and had no further encounters, then no 30-day data were reported. For RESP-LENS, ARI was defined as having one or more of the 130 ICD-10 diagnosis codes that was derived by consensus among the investigators and CDC personnel. Patients could be enrolled more than once, and unique patients with multiple visits were identified using their hashed medical record number.

Collected data are machine verified for completeness and formatting and then reviewed weekly by three CDC analysts for potential anomalies (e.g., unexpected changes in testing frequency). As needed, coding updates are made on a weekly basis to account for changes in test ordering patterns (e.g., use of new viral tests or updated codes).

### Laboratory Viral Testing

The decision to perform laboratory viral testing was at the discretion of the clinical care team at each hospital, or according to local policies. All sites required mandatory viral testing for all admitted patients up until approximately March of 2023, but this study only includes those patients with a respiratory complaint in the ED. All viral tests were performed on nasal or nasopharyngeal swabs with PCR or reverse transcription PCR. For each virus, each patient could be classified as either positive, negative, or not tested. All patient encounters were grouped into one of six mutually exclusive categories: i. COVID, ii. fluA, iii. RSV, iv. Multi-viral, v. Untested or vi. Negative test results.

### New onset VTE

In adherence with practice guidelines and to enable comparison with previous studies,^15,16^ VTE was defined as receiving a new ICD-10 code for either pulmonary embolism (26X) or deep vein thrombosis (87X) during the index visit or within the next 30 days. Patients with pre-existing ICD-10 codes of 26X or 87X, or “pulmonary embolism” or “deep vein thrombosis” documented in the structured past medical history list at the time of index visit were considered to have prior VTE.

### Statistical analysis

Frequencies and proportions were tabulated to describe categorical variables and quantiles or measures of central tendency were used to describe arithmetic variables, as appropriate.

Incidence rates with 95% confidence intervals (CI) were calculated to describe the number of new VTE cases per one-hundred ED patient encounters (%). Locally weighted regression was used to assess trends by profiling the weekly average new VTE incidence rate with 95% confidence limits. We also profiled the incidence rate of clinical tests that are routinely used to assess VTE risk (i.e., d-dimer blood test and computerized tomographic pulmonary angiography (CTPA) orders).

Log binomial regression models with generalized estimating equations were used to estimate incidence rate ratios with 95%CI, but also to test for interaction. Four sets of pre-specified models were fitted for each dependent variable. Model I was unadjusted and tested the general null hypothesis that there is no difference in the incidence (i.e., risk) of VTE among encounters with patients who either tested positive or negative for viruses or did not undergo viral testing. Model II adjusted for basic socio-demographics (i.e., age/race/sex/body mass index (BMI)/smoking category), whereas models III and IV adjusted for clinical histories of pulmonary embolism and deep vein thrombosis and model IV also included clinical comorbidities. Next, interaction terms were added to test if the risk of mortality within 30-days differed among encounters with patients who developed VTE depending on the results (or absence) of viral testing. Finally, conditional models were used to develop estimates for ED patients, inpatients, and patients that received intensive care.

Missing death information was interpreted as survival and missing ICU data was interpreted as ‘no exposure’; else, if data were unavailable in the EMR to definitively answer presence or absence of a variable, a “not available” code (e.g., ‘999’) was used and not included in tables or calculations.

Sensitivity analyses used multiple imputation to account for missing data by discriminant function and logistic regression for categorical variables or linear regression and predictive mean matching for continuous variables. We also used the Kaplan Meijer test for differences in survival time. Non-overlapping 95%CI are statistically different. The analysis was performed using SAS v9.4 (SAS Institute Inc., NC) and maps were produced using ArcGIS Pro 3.1.0 (ESRI Inc., CA) software.

## RESULTS

### Participant Characteristics

From 4,260,441 total ED visits, there were 620,303 (14.6%) encounters made by 496,620 unique patients at the 91 participating hospitals who received an ARI-qualifying ICD10 code. **Figure 1A** describes the demographic characteristics of the RESP-LENS cohort. Briefly, **63**% of the patients were below age fifty, **28**% identified as Black race, and about **half** were male. Body mass index (BMI) was recorded for only half of the participants; among them, **35**% were obese (BMI>30 Kg/m^2^).

Sixty-five percent of the cohort underwent viral testing; one of eight patient encounters tested positive for COVID (**13**%), which was fourfold more common than either fluA or RSV. Multi-viral test positivity was rare (<**1**%) and mostly involved a combination of fluA and RSV. Additional clinical characteristics of the full cohort, and of ED patients, inpatients, and the subset who required intensive care are described in **Figure 1B**.

### Incidence Rates and Trend

There were more than five thousand encounters with **3,483** unique patients who were met new VTE criteria (i.e., 0.7% [or **70** per 10,000] unique first patient encounters and 0.82% [or **82** per 10,000] total patient encounters; additional trend analyses are provided in the **Supplement**).

Figure 2A shows that the new VTE incidence rate was highest among encounters with patients whose viral test results were negative. By contrast, patients who tested positive for COVID, fluA or RSV developed VTE significantly less often. The weekly incidence rate ranged from **0.5%-to-2.0%** on average (**Fig 2C**); at no time during the period of surveillance was VTE incidence increased among ED encounters with patients who tested positive for viruses.

Blood D-dimer tests were ordered in nine percent of the encounters with patients who tested positive for COVID, which is higher than the six percent incidence rate among viral test negative patient encounters (***sFig 1A***). D-dimer orders among patients <18 years of age accounted for 1.8% of all D-dimer orders. In contrast to the pattern of D-dimer ordering, CPTA was ordered less frequently among encounters with patients who tested positive for each virus compared to viral test negative patient encounters (***sFig 1B***).

### Magnitudes of Association

From the entire sample, patients who tested positive for COVID, fluA, RSV, or a combination thereof, developed VTE less often than patients whose viral tests were negative. However, there were variations in the pattern of association by clinical setting. None of the viruses were associated with VTE among ED patient encounters (i.e., when the patient was not admitted) (***s***Fig 2), whereas inpatients who tested positive for each virus had significantly decreased VTE probability compared to their test negative peers (Fig 3). By contrast, among ICU patient encounters, fluA was associated with a significant threefold increased risk of VTE (***s***Fig 3).

### Mortality

The 30-day mortality rate was increased by nearly threefold among encounters with patients who developed VTE, adjusting for age, race and sex categories, plus clinical histories and comorbidities (Fig 4). This mortality association with VTE was observed in the full cohort, among non-admitted ED patients, and among all inpatients (***s***Fig 4). In contrast, viral test positivity was not associated with increased mortality in the entire cohort. Among inpatients requiring any ICU care, however, positive testing for COVID was associated with increased mortality risk, but not VTE, adjusting for patient characteristics and clinical histories/comorbidities (***s***Fig 5). There was no difference in the relationship between new VTE and death among patients grouped by the results (+/-) or absence of viral testing (interaction p > 0.5 among all encounters, inpatients and ICU encounters modeled separately).

### Sensitivity Analyses

Our findings were consistent when using discriminant function and logistic regression for binary/categorical variables or linear regression and predictive mean matching for continuous variables to account for missing data by multiple imputation (**Supplement Materials**). In addition to testing for differences in mortality risk, we also wanted to know if VTE patients who tested positive for viruses died sooner than their peers who did not test positive; we did not find evidence to support this possibility (Kaplan Meijer Logrank p= 0.87).

## DISCUSSION

In our study of nearly half a million patients who presented to U.S. emergency departments with respiratory complaints between January 2022 and June 2023, we found that laboratory confirmed positivity for SARS-CoV-2, influenza-A, RSV, or a combination thereof, is not associated with increased VTE risk. Additionally, we did not find evidence to support the possibility that viral test positivity modifies the association between VTE and heightened mortality risk. Our findings therefore do not support the inference that hospital patients with respiratory complaints who test positive for COVID in the ED setting should be considered at increased risk of VTE, nor that they will benefit from the administration of escalated- or full-dose prophylactic anticoagulation to prevent VTE.

### Comparison with previous studies

The VTE case-fatality rate in the RESP-LENS cohort is consistent with estimates from national samples and large PE registries.^17,18^ By contrast, previous observational studies tended to report higher VTE rates among patients who tested positive for COVID than was observed herein. For example, at least six metanalyses, that mostly describe a period of Hunan strain, predominance estimated that 7%-to- 32% of patients who tested positive for COVID develop VTE.^7-9,19-21^

While it is possible that differences in the underlying frequency distribution of viral subtypes played a role, selection bias is a more likely explanation for the difference between our findings and previous reports. Indeed, studies of high-risk patients who received intensive care are over-represented in the pooled meta-estimates, and in the RESP-LENS cohort the VTE incidence rate among COVID positive patients was increased by more than fivefold by restricting to ICU encounters (i.e., from 0.8% to 4.6%). Importantly, upwards of sixty thousand unique ED patients tested positive for COVID in RESP-LENS, which is more than are included in all previous meta-analyses combined.

The sum of our findings prompts us to infer that positive viral testing can serve as a viable alternative diagnosis for VTE in the emergency department setting. Namely, because, i) encounters with virus positive patients were less likely to be diagnosed with VTE, ii) among VTE patients, a positive viral test was not associated with heightened 30-day mortality risk, and iii) the reduced incidence of new VTE associated with a positive viral test does not appear to be the result of decreased screening based on the constant frequency distribution of D-dimer blood testing. Clinicians ordered CTPA testing more often in virus negative patients compared with virus positive patients. These observations raise the possibility of referral bias, whereby clinicians might have considered a positive viral test as an alternative diagnosis to PE and were more likely to order CTPA in virus negative patients.

### Strengths and weaknesses

The major strengths of this study are its prospective design and the large sample size afforded by the 91 hospitals participating in RESP-LENS that contributed data for all ten U.S. Department of Health and Human Services regions. RESP-LENS also uses a rigorously evaluated study protocol and a publicly available data management plan that involved a combination of automated and human monitoring of inbound data to assess quality during the ingestion process. This enabled us to account for the relatively frequent changes in coding, testing platforms, and policies across network sites.

Our study is limited by missing BMI data that was unavailable for about half the cohort; however, the results were consistent in sensitivity analyses with imputed data. Our findings are also specific to 2022 and part of 2023, and we do not have additional information about specific viral strains. We were also not able to incorporate information about immunizations or probe social determinants of health, but we plan to address these gaps in future work. It is also possible that some VTE cases may be misclassified (e.g., ICD-10 coding had 91% sensitivity and 99% specificity for the detection of PE in a previous study).^22^ Some 30-day outcomes might have additionally been missed, because we relied on the hospital system and their associated health exchange networks to identify post-discharge events. Lastly, as is the case with any observational study, ours is unable to distinguish between association and causation.

## CONCLUSION

Laboratory confirmed viral test positivity neither predisposes to heightened risk of new onset VTE nor modifies the 30-day mortality risk of affected patients in the RESP-LENS cohort of nearly half a million people who presented for emergency care.

## Data Availability

The RESP-LENS data management and data sharing plans are publicly available at https://dmptool.org/plans/70148.

## Contributors

SJK designed the statistical analysis plan, performed the analysis, and created figures other than the map and edited/approved this manuscript.

SJB created the map and edited/approved this manuscript.

CK assisted in weekly data transmission, regular participation in network meetings, and edited this manuscript.

DMC assisted in weekly data transmission, regular participation in network meetings, and edited this manuscript.

NK assisted in site-specific setup for data collection, weekly data transmission, regular participation in network meetings, and edited this manuscript.

CK assisted in site-specific setup for data collection, weekly data transmission, regular participation in network meetings, and edited this manuscript.

CAC co-conceived the work, assisted with funding, and provides technical advice for the conduct of the network and database content and edited the manuscript.

JAK conceived the work, obtained funding, organized the network and wrote the first draft of the manuscript.

## Acknowledgements

Funded by contract 75D30121C11813 and named the Respiratory Virus Laboratory Emergency Department Network Surveillance (RESP-LENS).

Site investigators involved in RESP-LENS effort (to be accessible as “collaborators” in PubMed):

| Principle Investigator | Institution (alphabetical) |
| --- | --- |
| Michael J. Roshon | Centura Health |
| Danielle E. Turner-Lawrence | Corewell Health William Beaumont University Hospital |
| Michael A. Puskarich | Hennepin Healthcare |
| Ka Ming Gordon Ngai | Icahn School of Medicine at Mount Sinai |
| Benton R. Hunter | Indiana University School of Medicine |
| Joseph Bledsoe | Intermountain Healthcare |
| James K. d'Ettiene | JPS Health Network |
| Stephen S. Lim | LCMC Health |
| Christopher W. Kabrhel | Massachusetts General Hospital |
| Esther J. Choo | Oregon Health & Science University |
| Steven MH Moore | Penn State Health |
| Katherine R. Buck | The Ohio State University Wexner Medical Center |
| David M. Beiser | The University of Chicago Department of Medicine |
| James G. Galbraith | The University of Mississippi Medical Center |
| Joby E. Thoppil | The University of Texas, Southwestern |
| Chris Kelly | The University of Utah |
| Edward W. Castillo | University of California, San Diego |
| Israel E. Green-Hopkins | University of California, San Francisco |
| Kristen S. Nordenholz | University of Colorado Anschutz Medical Campus |
| Faheem C. Guirgis | University of Florida Health |
| Bryan Wilson | University of Iowa |
| Michael L. Pulia | University of Wisconsin-Madison |
| Stacey House | Washington University School of Medicine in St. Louis |
| Justine M. Pagenhardt | West Virginia University |

---

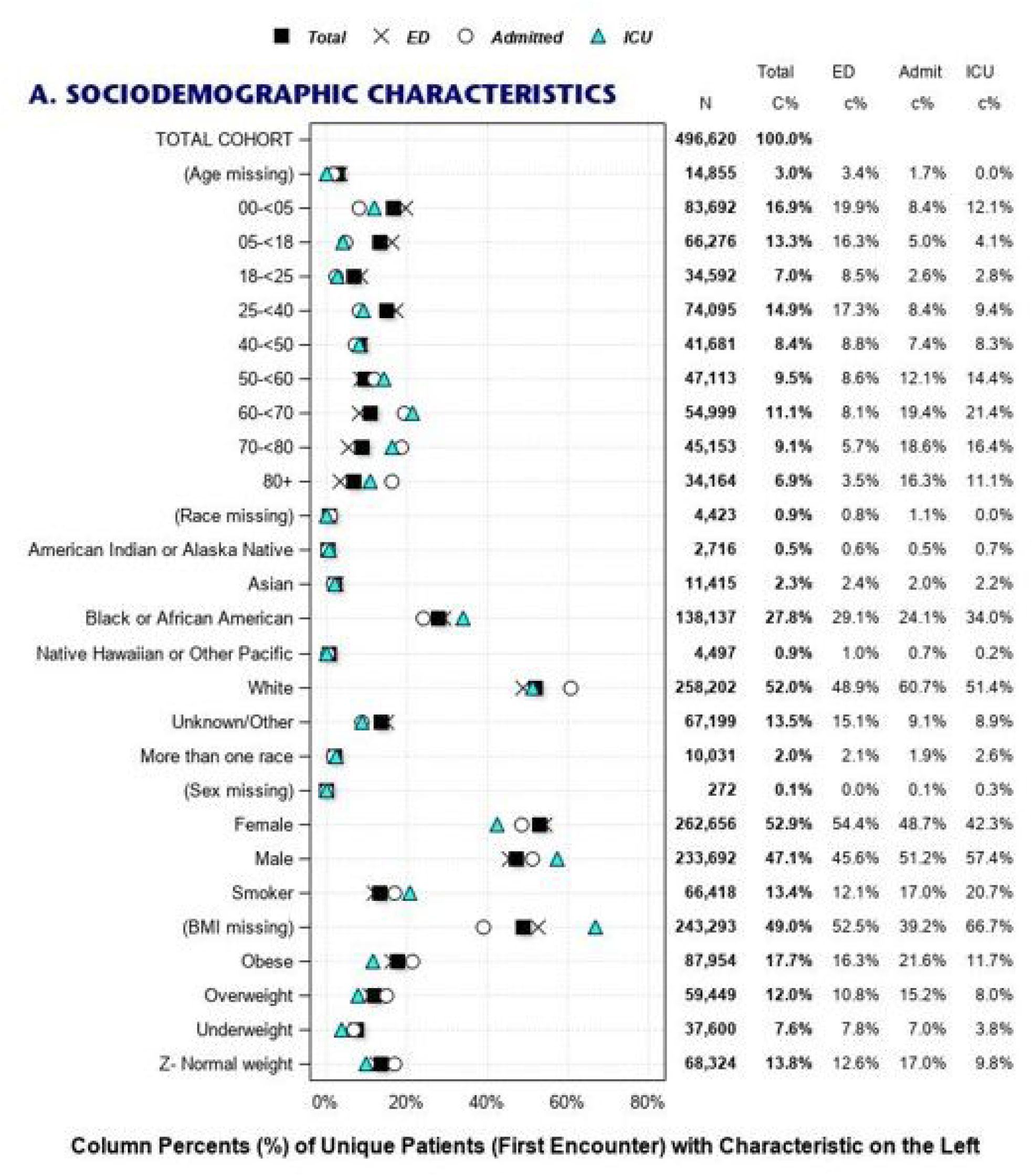

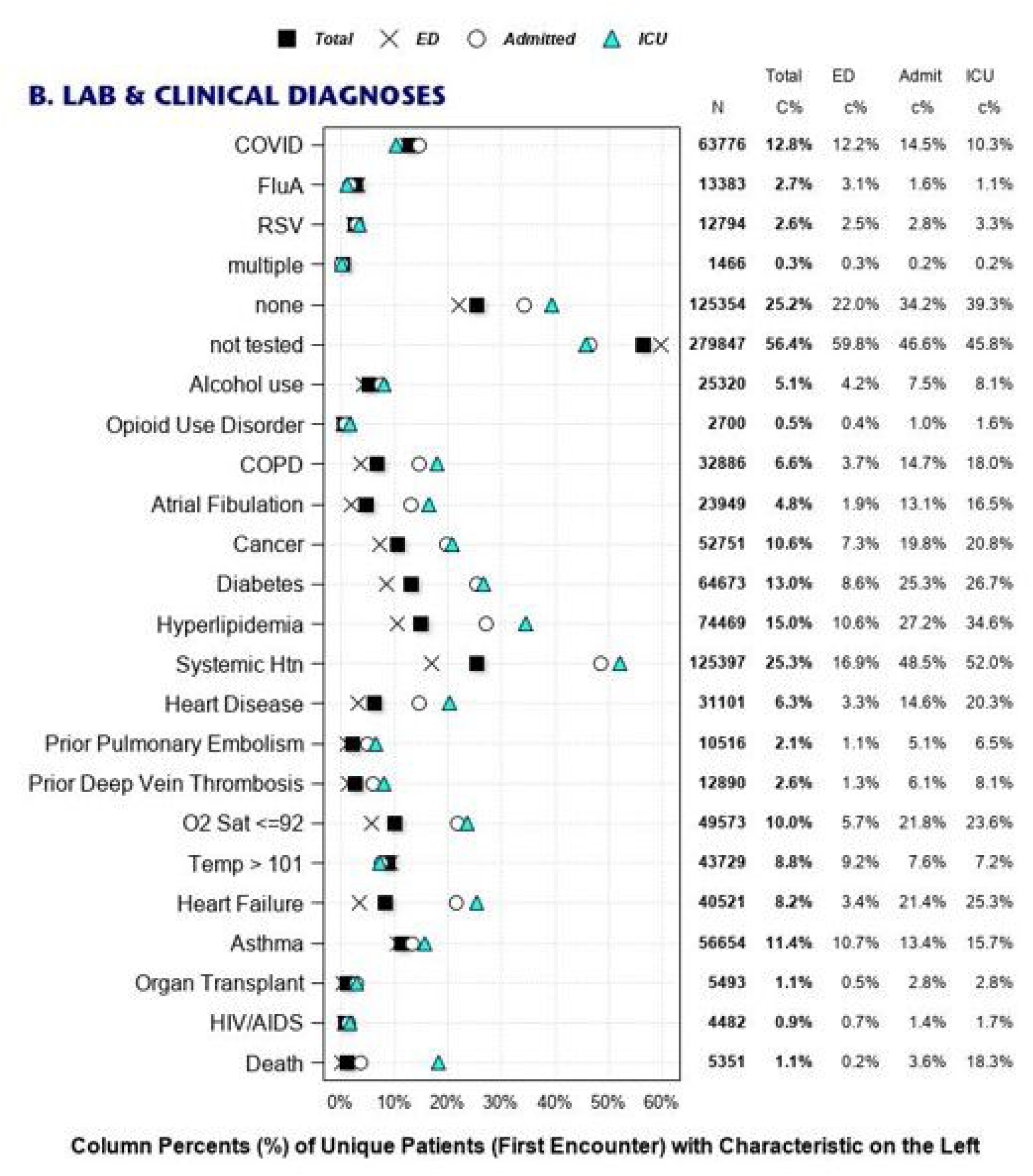

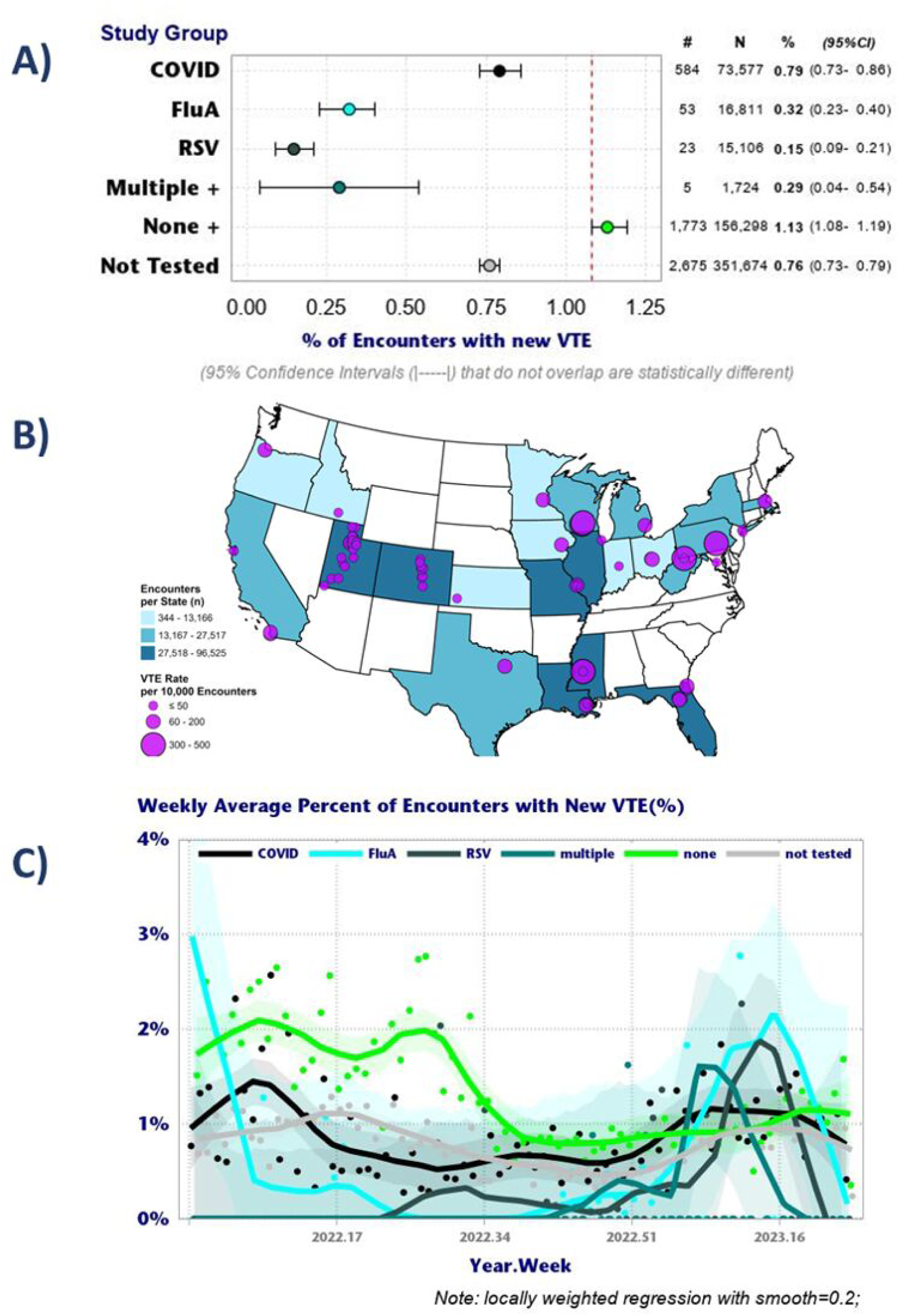

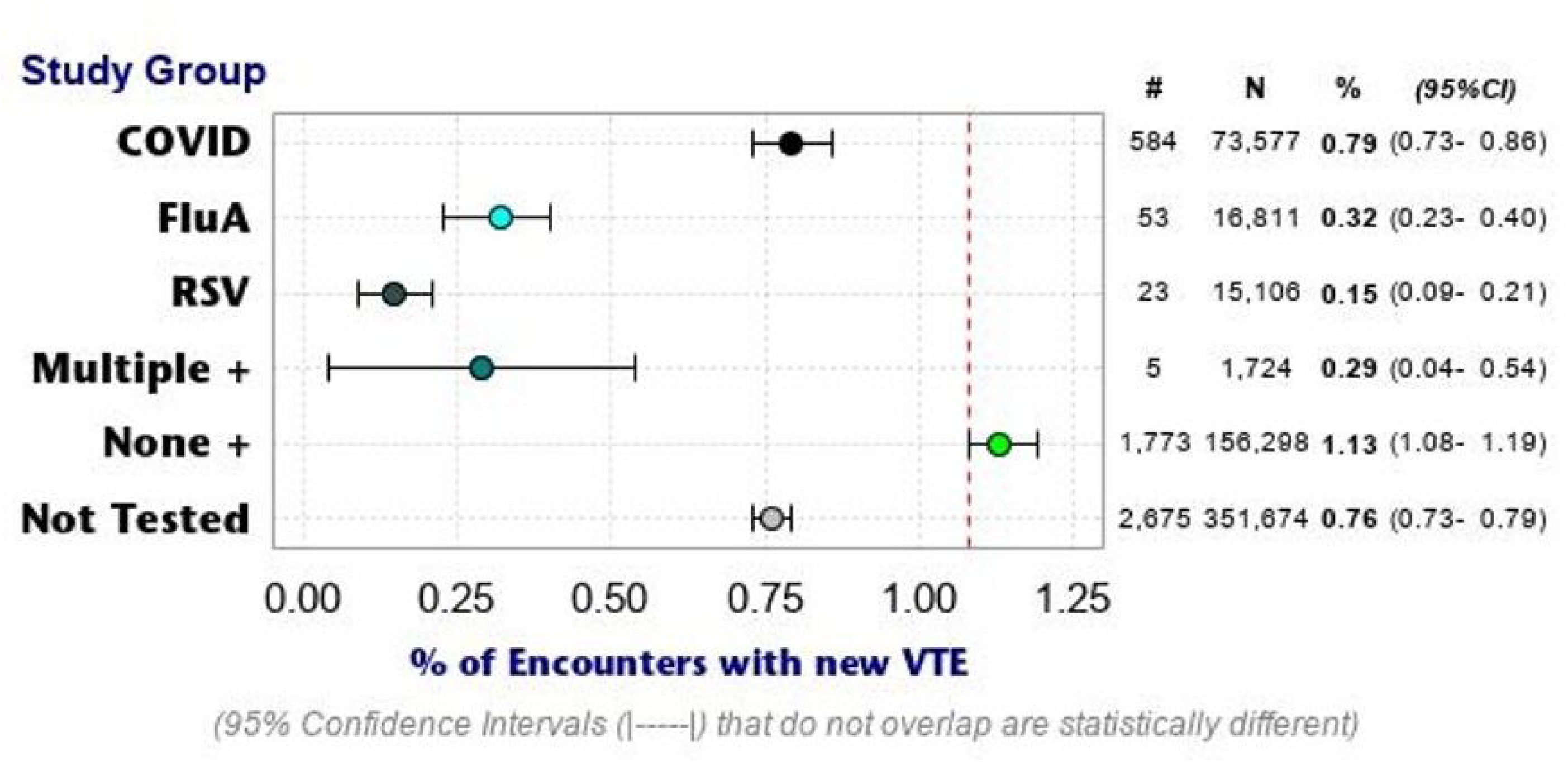

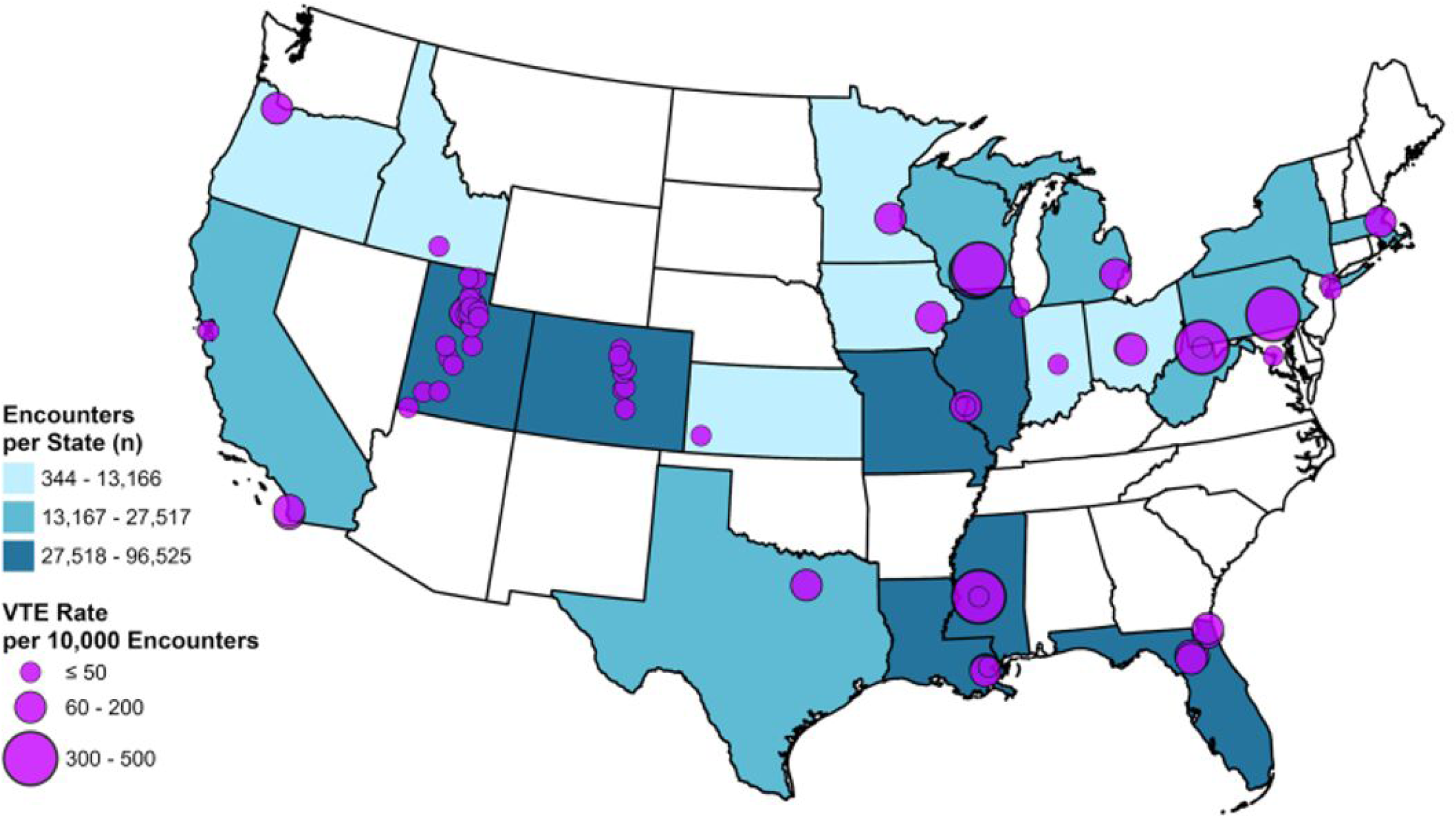

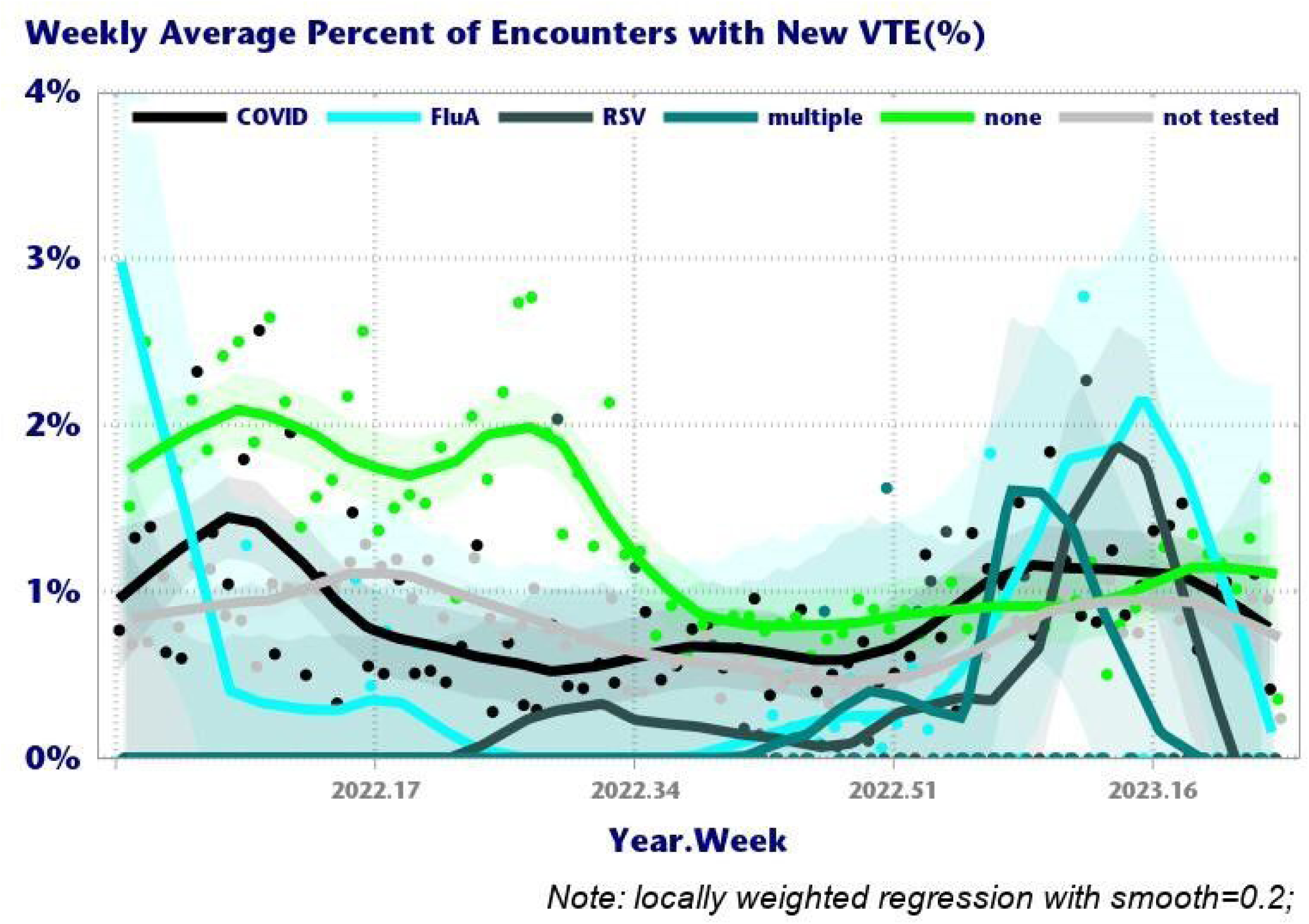

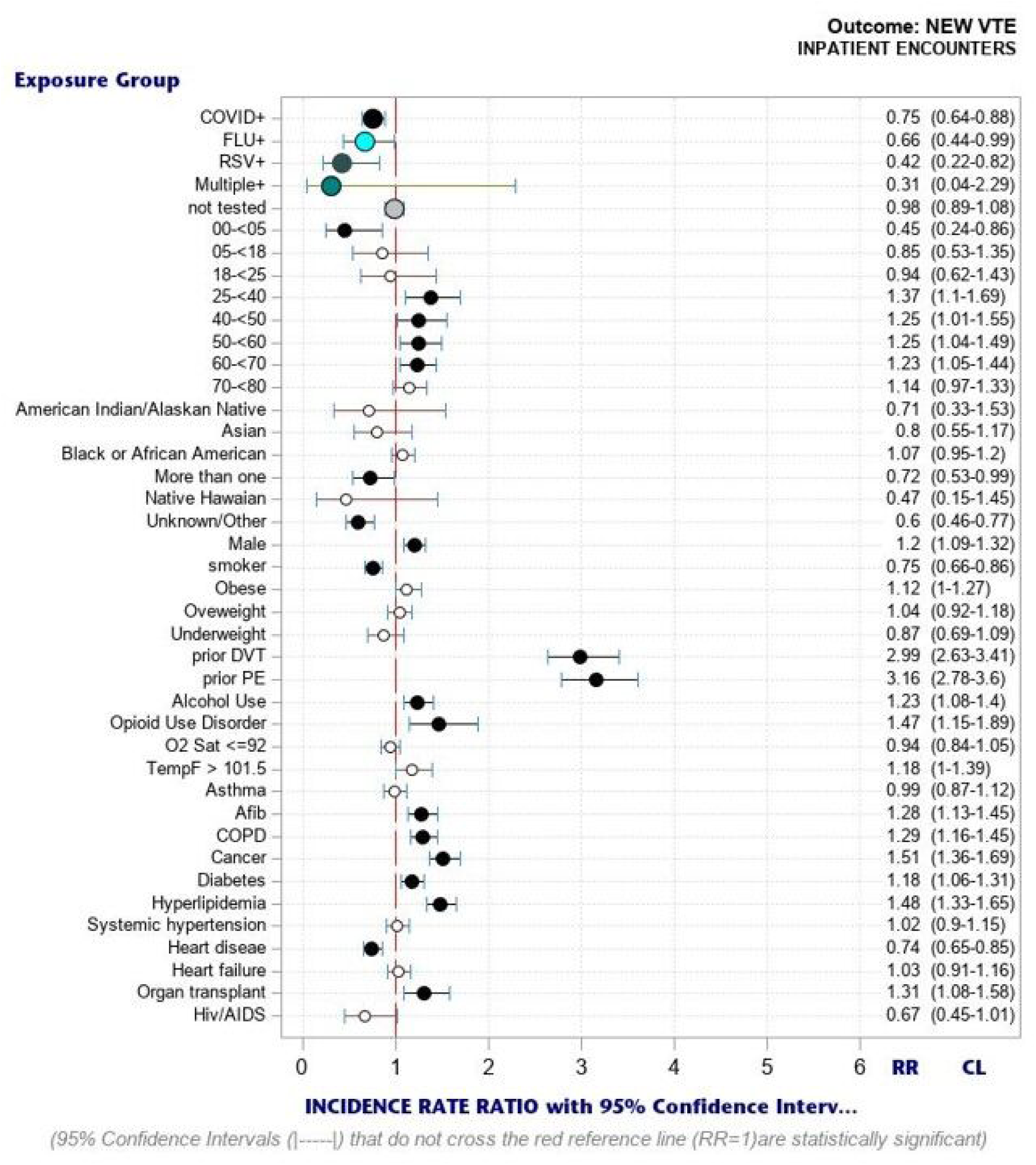

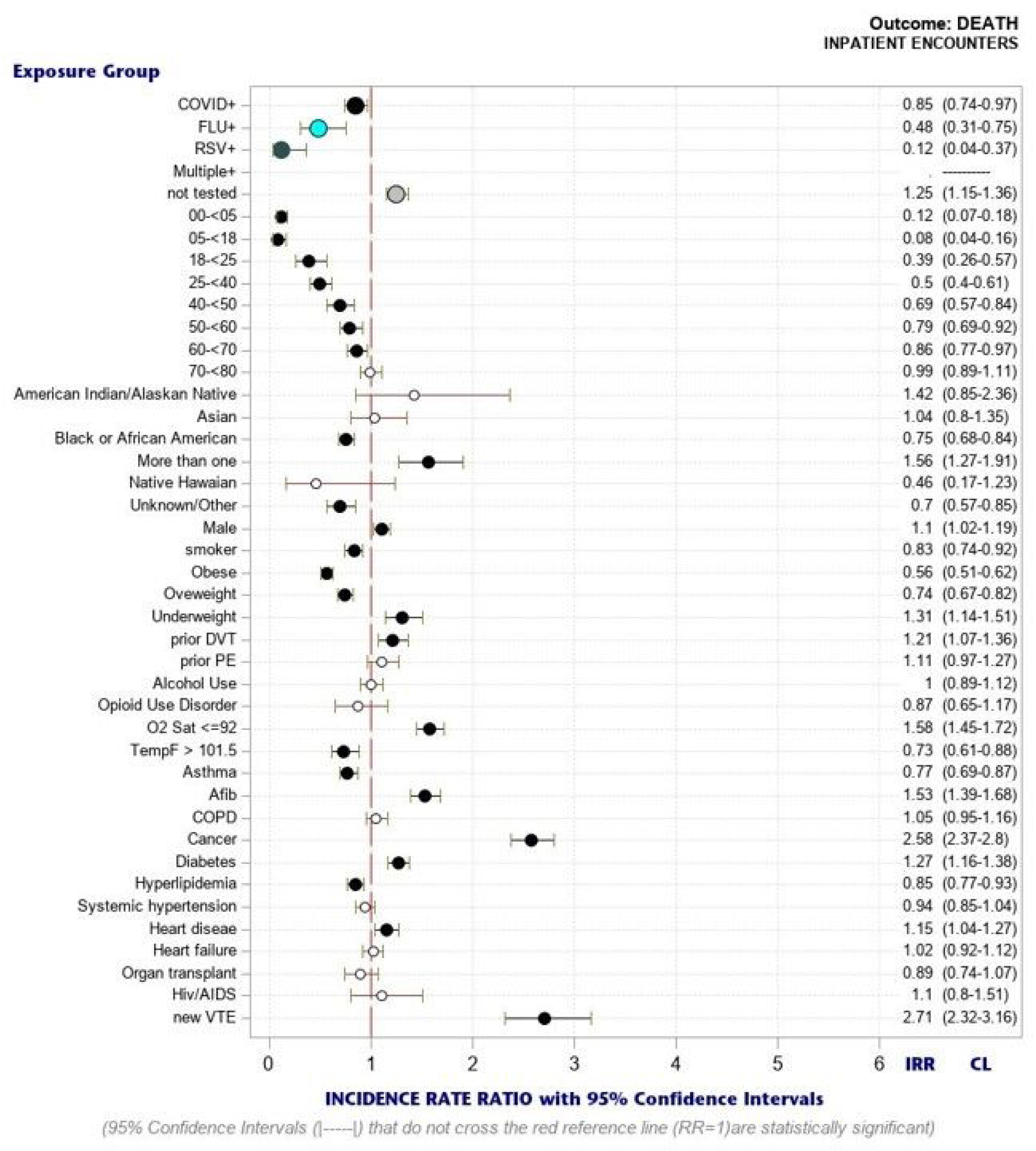

